# Plasma proteomic profiling identifies country-specific diversity in visceral leishmaniasis patients across East Africa

**DOI:** 10.64898/2026.10.16.26365713

**Authors:** Joao Cunha, Ayenew Addisu, Daniel Matano, Brima M. Younis, Alice Bayiyana, Rajiv Kumar, Rahul Tiwari, Jessica Engel, Karen Hogg, Rebecca Wiggins, Wilson Biwott, Finnley Osuna, Christine Ichugu, Ayalew Jejaw Zeleke, Eleni Ayele, Obondo James Sande, Eltahir A.G. Khalil, Hussam M.H. Ibrahim, Mahmoud A. Mahmoud, Ahmed I.B. Zakaria, Brenda Adiko, The Immstat@Cure Consortium^, Peter O’Toole, Flavia D’Alessio, Charles J.N. Lacey, Christian R. Engwerda, Shyam Sundar, Jane Mbui, Asrat M. Hailu, Margaret Mbuchi, Ahmed M Musa, Joseph Olobo, Paul M. Kaye

**Affiliations:** York Biomedical Research Institute, Hull York Medical School, University of York, York, UK; Department of Medical Parasitology, School of Biomedical and Laboratory Sciences, College of Medicine and Health Sciences, University of Gondar, Ethiopia; Center for Clinical Research, Kenya Medical Research Institute, Nairobi, Kenya; Institute of Endemic Diseases, Khartoum, Sudan; Department of Immunology and Molecular Biology, Makerere University, Kampala, Uganda; Centre of Experimental Medicine & Surgery, Institute of Medical Sciences, Varanasi Hindu University, Varanasi-221005, India; QIMR Berghofer Medical Research Institute, Brisbane, Australia; BioSciences Technology Facility, Dept of Biology, University of York, York, UK; Leishmania Research and Treatment Center, University of Gondar Comprehensive and Specialized Hospital, Gondar, Ethiopia; European Vaccine Initiative, Heidelberg Germany; Department of Medicine Institute of Medical Sciences, Varanasi Hindu University, Varanasi-221005, India

**Keywords:** plasma proteomics, visceral leishmaniasis, kala azar, treatment response, Olink

## Abstract

**Background:** Visceral leishmaniasis (VL) is the most severe form of leishmaniasis and is caused by parasites from the *Leishmania donovani* complex. VL is endemic in 54 countries, with East Africa (EA) accounting for ∼70% of the global burden. Therefore, there is a pressing need to prioritize VL research in EA. Transcriptomics analysis in rodent models and patients following treatment have shown that immune homeostasis is not restored by the end of treatment. However, comprehensive proteomic studies in VL remain scarce, and the few available studies were conducted outside EA.

**Methodology/Principal Findings:** As a part of ImmStat@cure consortium, we used the Olink Reveal proximity extension assay to profile circulating proteins in VL patients from Ethiopia, Kenya and Sudan. By comparing patients at admission and 17 days after treatment with SSG-PM, we have identified a systemic Th1-cytotoxic inflammatory profile at admission, characterized by high abundance of IFNγ, IL-1B, IL-6, and the granzymes A and B. Chemokines with high abundance included CXCL8/10/11, CCL3, CCL4, CCL8, CXCL8 and CCL20, consistent with the recruitment of monocytes, macrophages, T cells, and NK cells. There were also evidence of immune regulation/exhaustion, with high levels of PD1, PD-L2, IDO1 and KYNU. Although the immune profiles were similar at admission, country specific differences were observed after treatment. Kenyan patients exhibited a sustained inflammatory profile and signatures of myeloid recruitment, characterized by high levels of IL-1B, IL-6 , IL-12A, CXCL8, CCL2, CCL3, CCL4 ,CCL7 and CEBPB. Sudanese patients showed signatures associated with lymphocyte activation and immune regulation.

**Conclusions/Significance:** These results highlight the complexity of the immune changes associated with VL, identifying both core conserved profiles at admission and country-specific profiles after treatment. The data provided here will provide a valuable resource for future comparative studies across more diverse different geographic regions, for linkage studies that assess the impact of parasite genotype on immune / metabolic response and for use during the evaluation of new therapies against VL.

## Introduction

Visceral leishmaniasis (VL) is the most severe form of leishmaniasis, characterized by splenomegaly, hepatomegaly, weight loss, pancytopenia and hypergammaglobulinemia. It is primarily caused by parasites of the *Leishmania donovani* complex, which are transmitted by infected sand fly vectors [1]. As of January 2026, VL is endemic in 54 countries, where ∼85% of the global cases were reported in seven countries: Brazil, Ethiopia, India, Kenya, South Sudan and Sudan [2]. With the control of VL in India, Nepal and Bangladesh [3, 4], East Africa currently bears ∼70% of the global VL burden, with fatality rates ranging from 4-11% [5, 6]. Given this significant epidemiological burden, there is a pressing need to prioritize research and control efforts in East Africa.

The immune response in patients with VL before and after treatment has been extensively investigated using transcriptomic analyses. In rodent models of VL, transcriptomic profiling following drug treatment has shown that systemic immune, metabolic, and tissue remodelling abnormalities are not fully restored to homeostasis by the end of treatment (EOT) [7, 8]. RNA sequencing of peripheral blood from Brazilian patients revealed widespread alterations in interferon signalling, chemokine and Toll-like receptor pathways and neutrophil-associated responses, highlighting immune changes associated with infection and clinical recovery [9]. Similarly, whole-blood microarray analysis of patients with VL in India identified *IFNG* as the principal hub gene in active VL. Comparisons between active and treated cases further revealed enrichment of transcripts for interleukin signalling (*IL1*, *IL3*, *IL4*, *IL6*, *IL7* and *IL8*) and IFNγ-inducible chemokines (*CXCL9*, *CXCL10*, and *CXCL11*) [10]. A longitudinal transcriptomic profiling of African patients with VL and HIV coinfection, including individuals who were cured or had treatment failure, identified a four-gene signature (*PRSS33*, *IL10*, *SLFN14*, and *HRH4*) with potential to discriminate treatment outcomes [11]. Taken together, these studies highlight the importance of Th1 immune activation in VL, where the expression levels of relevant immune mediators only partially revert back to homeostasis at the end of treatment.

The transcriptomic findings are broadly supported by studies investigating systemic protein changes during VL treatment, which have been limited to single-country cohorts or have relied on limited cytokine or protein marker panels. These studies consistently identified IFNγ, IL-10, and TNF as the principal immune mediators associated with active disease in several countries, with more limited evidence supporting a role for Th17-related responses [12–15]. On the other hand, comprehensive proteomic studies in human VL patients remain scarce, where the few studies available have been conducted outside East Africa. Torres *et al.* analyzed the proteomic composition of circulating extracellular vesicles from Spanish VL patients with active disease and at 1, 3 and 6 months following treatment with liposomal amphotericin B. Using LC–MS/MS on pooled peripheral blood samples, they reported a reduction in immune-related pathways after treatment, including proteins associated with type I interferon, TNF and IL-17 signalling [16]. Two studies based on comparative proteomic analyses of samples from Indian VL patients and healthy controls using two-dimensional electrophoresis (2DE) based methods, identified differential expression of proteins including α1-antitrypsin (A1AT), α1B-glycoprotein (A1BG) and serum amyloid A1 (SAA1) and C1 inhibitor [17, 18]. Despite these advances, our understanding of systemic protein alterations in VL and their relationship to treatment outcomes remains limited, particularly in East Africa.

Recent advances in high-dimensional plasma proteomics provide an opportunity to define systemic host responses to infection at substantially greater depth than previously possible. Proximity extension assay (PEA) proteomics has emerged as a highly sensitive and reproducible approach for high-throughput protein profiling from small sample volumes [19], and can quantify low-abundance proteins that are usually not detected by traditional MS approaches [20]. Olink Reveal is an NGS-based implementation of PEA that enables the relative quantification of 1,034 predefined proteins, spanning representatives from the Top level Reactome pathways, including 537 immune-related proteins. Because of its high analytical precision, low sample volume requirements, and standardized workflow, PEA is well suited for large multicentre studies where reproducibility across sites and batches is essential. Here, as a part of ImmStat@cure [23], we used the Olink Reveal proximity extension assay platform to profile more than 1,000 circulating proteins in patients with VL from Ethiopia, Kenya and Sudan at admission and at 17 days after treatment (end of treatment; EOT) with SSG-PM, and compared with healthy controls. By comparing proteomic profiles across countries, disease states and treatment, we sought to determine whether VL is associated with conserved systemic signatures across East Africa or whether substantial country-specific heterogeneity exists. This study provides the first large-scale multi-country proteomic analysis of VL in East Africa and identifies both shared and geographically distinct host responses associated with active disease and treatment.

## Methods

### Ethical and regulatory approval

This multicentre clinical study was approved by ethical and regulatory agencies in the UK (Department of Biology Ethics Committee reference PK202001, Hull York Medical School Ethics Committee reference 22-23.58), Ethiopia (University of Gondar Ethics Committee reference V/P/RCS/05/495/2019; Ministry of Education reference 7/2–324/M259/35), Kenya (National Commission for Science, Technology and Innovation references NACOSTI/P/21/11096/P6958/25/415974; KEMRI Scientific and Ethics Review Unit reference KEMRI/SERU/CCR/0169/4113), Sudan (University of Khartoum Ethics Committee reference FM/DO/EC; Sudan Ministry of Health approval 7-12-20) and India (Dean/2023/EC/6561 dated 09.10.2023 and IMS/IEC/June/2026/459 dated 22/06/2026). All studies were conducted in accord with the Declaration of Helsinki and with participant informed consent. Formal written consent to participate in the study was obtained from the patients or their legal guardians. The study did not involve modification of existing standards of care provided to patients.

### Study sites and patient recruitment

This work was conducted as part of Immstat@cure, an observational multicentre clinical study to assess the clinical and immunological status of VL patients before and at the End of Treatment (EoT) with SSG/PM/MG, across four countries in EA: Ethiopia, Kenya, Sudan and Uganda. In the current project, only samples from Ethiopia, Kenya and Sudan were analysed, based on sample availability and quality. First and last recruitment for each country were: Ethiopia, 01/2024 and 06/2024; Kenya, 11/2022 and 04/2024; Sudan, 01/2023 and 05/2024. The study was prospectively registered on ClinicalTrials.gov (ClinicalTrials.gov ID: NCT04342715).

The project details, including recruitment protocol, inclusion and exclusion criteria are provided elsewhere [23]. In short, Patients were recruited by active case detection and self-attendance at study hospitals. Ethiopian patients and endemic controls were recruited from the Gondar region and processed at the University of Gondar (UoG). In Kenya, patient recruitment was conducted in the Chemolingot Sub-County Hospital in Baringo county, and healthy volunteers were recruited from the Center for Clinical Research, Nairobi County. Samples were processed at the Kenya Medical Research Institute (KEMRI). In Sudan, patient and endemic controls recruitment were conducted at Soba Hospital, Khartoum, the Institute for Endemic Diseases (IEND), the El Hassan Centre for Tropical Medicine, Doka, Gedarif State and the University of Khartoum, Sudan. As a further comparator group, samples were obtained from Indian participants recruited from the Kala-Azar Medical Research Center (KAMRC) in Muzaffarpur, Bihar, India.

VL diagnosis was performed using the rK39 rapid diagnostic test [24], and confirmed by amastigote identification through microscopic examination of aspirates (spleen, bone marrow or lymph nodes) stained with Giemsa and observed at 10x eyepiece and x100 magnification. Patients in EA were treated with SSG (20mg/Kg/day; i.m.) and PM (15 mg/kg/day; i.m.) for 17 days. Clinical, haematological and immunological data, as well as peripheral blood samples for proteomics were collected on admission and at the end of treatment. The occurrence of relapse and PKDL was assessed at 6 months follow up after completing treatment. Patients in India were treated with Ambisome (single dose, 10 mg/kg body weight) and assessed for treatment response at day 30.

### Quality control/Filtering and data scaling

Patient and healthy volunteer plasma samples from all countries were randomized across plates prior to protein profiling using the Olink Reveal platform by Proximity extension assay (PEA) (Olink Proteomics, Uppsala, Sweden). In PEA, pairs of antibodies labelled with complementary DNA oligonucleotides bind to their target protein. When both antibodies bind in close proximity, the oligonucleotides hybridize and are extended by DNA polymerase to generate a unique DNA barcode, which is quantified by next-generation sequencing (NGS). This strategy provides high specificity while enabling multiplexed measurement of hundreds to thousands of proteins in parallel. The analytical performance of Olink Reveal has been recently validated using recombinant protein spike-in experiments, demonstrating excellent linearity across the assay’s dynamic range (R² = 0.922–0.953) [21]. Protein relative abundance was reported as Normalized Protein eXpression (NPX), Olink’s log2-scaled values, normalized using internal controls that accounts for technical and plate variation across runs.

Several quality control steps were used for data cleaning. The Limit of Detection (LOD) for each protein was estimated using the function “olink_lod” from the Olink Analyze R library (https://cran.rstudio.com/web/packages/OlinkAnalyze/vignettes/OlinkAnalyze-Vignette.html) and the model data “Reveal_Fixed_LOD_24_04_2026”, obtained from the Olink repository. The model data is supplied as **Supplementary_Model**, for reproducibility. Only samples that passed Olink sample quality control (SampleQC = PASS) and had at least 400 of the 1034 proteins detected above LOD were used in downstream analysis. Protein assays with NPX values below LOD in more than 90% of the samples from all datasets were removed, resulting in 1018 proteins being used in downstream analysis. For retained proteins, NPX values below LOD were imputed as the minimum observed NPX among all patients for that protein minus 0.01 NPX units.

To minimize residual variation arising from differences in sample collection, handling, storage, shipping or processing across countries that may not be fully addressed by Olink normalization procedures, NPX values from each country were scaled separately, by subtracting the mean NPX of healthy volunteers in the country. As NPX is in log_2_ scale, this is equivalent to dividing by the HV mean in linear scale.

Samples from India were processed and scaled separately from samples from Ethiopia, Sudan and Kenya. Hence, direct comparisons of protein levels between India and the East African countries were not performed.

### Statistical analysis

All comparative statistical analysis and data representation were generated in R (version). All scripts can be obtained from the GitHub repository (https://github.com/jaumlrc/Immstat_cure_OlinkProteomics).

### Evaluating the clinical data

Differences in the clinical data from VL patients pre and post treatment were assessed using non-parametric paired Wilcoxon signed rank test, while comparisons between VL patients and convenience controls were performed using the Wilcoxon rank sum test (equivalent to Mann-Whitney test). P-values were adjusted for multiple comparisons using Benjamini-Hochberg (BH) false discovery rate (FDR) method.

### Dimensionality Reduction and Differential Expression Analysis

Dimensionality reduction and visualization was generated by principal component analysis (PCA) and Uniform Manifold Approximation and Projection (UMAP), based on centred and scaled NPX, using respectively the functions prcomp and umap [25]. UMAPs were generated using the following parameters: min_dist = 0.1, metric = “euclidean”, n_epochs = 1000, n_neighbour = 15, seed = 1234. Graphical representations were generated using ggplot and factoextra. Heatmaps were generated using the pheatmap package, with hierarchical clustering of samples and proteins by Manhattan distance and unweighted pair-group method with arithmetic mean (UPGMA; average linkage).

Diffusion maps and trajectory inference were generated using a combination of the diffusionMap, mclust and slingshot R packages. First, the diffusion maps were generated using the *diffuse* function from the diffusionMap package, based on a Euclidean distance matrix calculated from NPX values across patients. Patient clusters were defined by Gaussian mixture modeling (GMM) using the *Mclust* function from the mclust package, based on the first two diffusion map components (DC1 and DC2). Then, the trajectory inference was generated using the slingshot package, with fitted lineage curves in the two-dimensional diffusion map embedding using the Mclust derived clusters as input.

Differential protein expressions were assessed by linear regression using the R package *Limma*. Comparisons between groups were performed using linear models (lmFit function) with empirical Bayes moderation (eBayes), adjusting for Age covariate and with Benjamini-Hochberg (BH) false discovery rate (FDR) multiple test correction [26]. Results not adjusted by age are reported as supplementary files, for comparison. For comparisons between V1 and V2 samples, patient identity was included in the design matrix to account for the paired study design. As the V1–V2 comparison was performed longitudinally using samples collected from the same individuals at two time points, age adjustment was not required. As NPX values are reported on a log_2_ scale, differences in NPX between groups are equivalent to log_2_ fold changes (log2FC). Therefore, differences in NPX are reported as log2FC. Proteins were considered significantly differentially expressed if they met an adjusted *p*-value < 0.05 and a log2FC > 1 for inter-stage comparisons, or an adjusted *p*-value < 0.05 and an absolute log2FC > 0.5 for comparisons between countries within the same stage. Pearson correlation between log2FC values was estimated using the cor.test function, in R.

Protein–protein interaction network analysis was performed using the STRING database web function (https://string-db.org/), with lists of differentially expressed proteins. The following active interaction sources were included: Experiments, Gene Fusion, Databases, Co-occurrence and Co-expression. A medium confidence score (0.4) was used as a minimum required interaction score. Network clusters were identified using k-means clustering, implemented in STRING. Only proteins that were present in a network were included in the final visualisations.

Over Representation Analysis (ORA) for each set of overexpressed proteins was performed using the ClusterProfiler R library [27]. Enrichment analyses were conducted for Gene Ontology (GO), Kyoto Encyclopedia of Genes and Genomes (KEGG) pathways, and Reactome pathways. The background universe was restricted to the 1,018 relevant proteins in the Olink Reveal panel. Statistical significance was determined using Benjamini–Hochberg (BH) false discovery rate (FDR), with an adjusted P-value < 0.05

### Custom module score and Transcription Factor relevance analysis

The custom module score analysis is described in detail in **Fig. S13**. NPX values for each protein were centred and scaled using all samples. This standardization ensured that each protein contributed equally to the module score, such that the score reflected relative variation in protein abundance rather than differences in the magnitude of NPX values. For each module in each sample, the module score was calculated as the mean of the standardized NPX values of all proteins assigned to the module in that patient. Module scores were compared between groups (e.g., V1 versus HV) using linear models (lmFit function) implemented in the limma R package, with empirical Bayes moderation (eBayes) and age correction [26]. For comparisons between V1 and V2 samples, patient identity was included in the design matrix to account for the paired study design, and no age adjustment was needed.

The assessment of master regulator transcription factors that vary between disease stages was done using decoupleR [28], and DoRothEa regulons [29]. Differential expression t-statistics obtained from each comparison, (e.g., pre-treatment versus healthy volunteers) were used as an input to the Univariate Linear Model (ULM) method implemented in decoupleR. This analysis was performed with the highest confidence DoRothEA regulons (confidence levels A and B) as the prior regulatory network. P-values were adjusted for multiple testing using BH, and the transcription factors with an adjusted p-value < 0.05 were considered significantly differentially active.

### Linear modelling of protein–clinical trait associations

Associations between protein expression and clinical traits were analysed using the Limma R package. Only patients with complete NPX and clinical data at both pre– and post–treatment timepoints were included. Protein NPX values were modelled as a function of each clinical trait and disease stage (pre vs. post–treatment), with country of origin and age included as covariates. To account for within–participant repeated measurements, duplicateCorrelation was applied with patient ID specified as the blocking factor. Variance estimates were stabilised using empirical Bayes moderation. Contrasts were defined to estimate the association between each clinical trait and protein expression at V1 and/or V2. P–values were adjusted for multiple testing using the Benjamini–Hochberg false discovery rate (FDR), and associations with FDR–adjusted p–values < 0.05 were considered statistically significant.

## Results

### Olink Reveal proteomics identifies heterogeneity in VL patients from East Africa

To determine the plasma proteome of 87 VL patients at presentation (V1) and end of treatment (EOT; V2) and 71 healthy convenience controls (HV) recruited from Ethiopia, Kenya and Sudan, we used the Olink Reveal platform (1018 proteins passing QC criteria; see Methods). Following QC and filtering, 54 VL patients and 64 HV met the inclusion criteria for downstream analysis (29, 15 and 10 patients, and 27, 27 and 10 HV from Ethiopia, Kenya and Sudan, respectively; **Fig. S1**). Clinical, biochemical and haematological data summaries are provided in **Fig.S2**, and **Fig. S3**. NPX values were rescaled based on the mean NPX value of HV in each country, to remove potential confounding bias from differential sample handling across countries (**Fig. S4**).

Applying the uniform manifold and projection (UMAP) technique, we observed clear separation of HV from patients at V1, though with country specific differences in the degree of separation (**Fig. 1A**). As anticipated from previous studies in murine models [7, 30], and in our preliminary analysis of this patient cohort using a panel of inflammation markers [23], EOT samples localised to an intermediate position in the UMAP projections and heatmap of NPX values (**Fig. 1A, B, Fig. S5**). A Diffusion map based on the Euclidean distances of the NPX profiles across patients suggested that patients from Ethiopia and Sudan, but not Kenya, shared a similar biological trajectory, with EOT samples occupying an intermediate position between V1 and HV (**Fig. 1C**). An elbow plot of diffusion components indicated a clear inflection point between DC1 and DC2 (**Fig. 1D**). The distribution of patients along DC1 confirmed the similarity between Ethiopian and Sudanese samples whereas samples from Kenya were distinguished along DC2. Fitting a Gaussian mixture model (GMM) resolved the data into discrete clusters and Slingshot trajectory inference projected onto these clusters identified two distinct lineage structures, with samples from Kenya following a different inferred trajectory compared to those from Ethiopia and Sudan (**Fig. 1E**). Finally, we quantified distribution across trajectories, confirming enrichment of trajectory 2 in Kenyan patients (Chi-squared p-value 6.86E-24, residuals in Lineage 2: 10.29) (**Fig. 1F**). In summary, the analysis of the plasma proteome suggests country-based heterogeneity in VL in East Africa, separating Kenyan patients from those in Ethiopia and Sudan.

**Figure 1:**
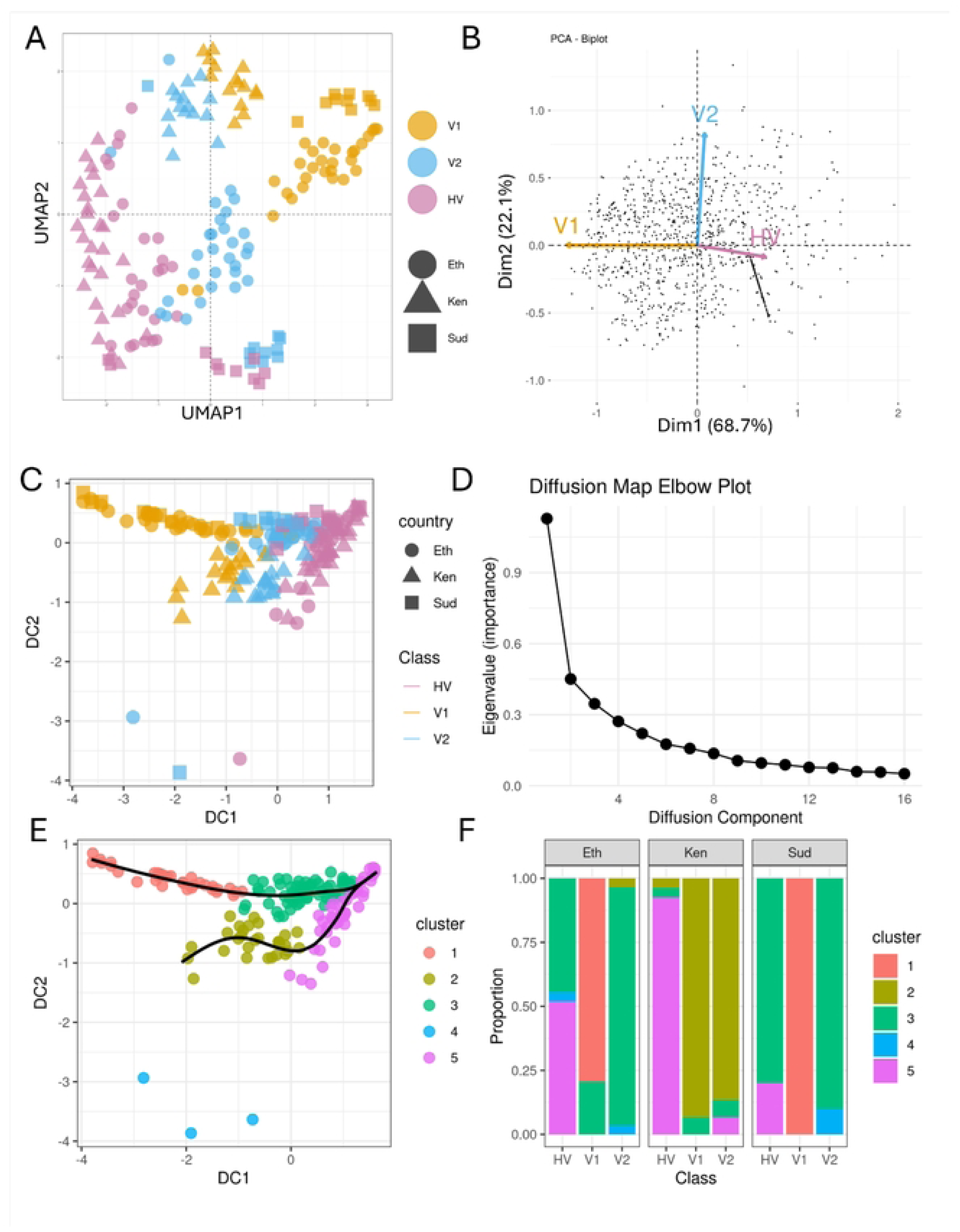
Treatment response trajectories in East African VL patients. **A)** UMAP of centred and scaled NPX values across all countries and disease stages. Each point represents a patient, where disease stage and country of origin are represented respectively by colours and shapes. **B)** Inverted PCA biplot illustrating the trajectory of inflammation resolution and homeostasis restoration. Arrows represent disease stages, where the angle between arrows reflects the similarity in their proteomic profiles. PC1 represents ∼70% of the variance and separates V1 (pre-treatment) from HV (healthy volunteers); while V2 (post-treatment) is positioned between both, consistent with partial recovery. PC2 represents ∼20% of the variance, and is primarily influenced by V2, with a smaller contribution of HV. Each point corresponds to an individual protein. **C)** Diffusion map of patient proteomic profiles based on Euclidean distances from protein NPX values. Each point represents a patient, and the X and Y axis represent, respectively, the DC1 and DC2. Points are coloured by disease stage (HV, V1, V2) and the countries are represented by shapes. **D)** Eigenvalues for each diffusion component (DC). Components with higher eigenvalues represent more of the intrinsic distribution of NPX in the proteomic space. **E)** Diffusion map as in **C**. Colours represent the clusters, identified by Gaussian mixture modelling of DC1 and DC2, and represented by colours. The black line illustrates the Slingshot-inferred reconstructed trajectory structure (after omission of the outliers forming cluster 4). **F)** Proportion of samples from each country and disease stage that corresponded to each cluster.

### VL patients share a similar core Th1 inflammatory profile at admission

To identify potentially common proteomic signatures associated with VL and explore the basis for the country-specific differences observed above, we analysed proteins that were differentially expressed between V1 and HV. Age correction was applied to all comparisons to account for differences in age distribution among patients, controls, and countries (with impact of age correction provided in **Fig. S6)**. Changes in protein abundance were consistent across countries, with very strong Pearson proteome-wide correlations of log2FC values (∼0.6-0.8) (**Fig. S7**). A total of 533 proteins (250-434 proteins in each country) were more abundant at V1, compared to only 12-21 proteins were more abundant in HV (**Fig. S6**, **Fig. S8**, **Table S1**). This imbalance likely reflects the enrichment of immune-related proteins in the Olink Reveal panel, and the strong systemic immune activation observed in VL.

A total of 176/533 (33%) proteins were consistently more abundant in V1 across all countries (**Table S2**), while another 142/533 (27%) were common to Ethiopia and Sudan (**Fig.2A**). STRING network analysis of the 176 shared proteins revealed highly interconnected immune networks, consistent with a strong Th1/cytotoxic inflammatory response, including IFNγ, IL-1B, IL-6, and the granzymes GZMA and B. Strongly induced chemokines included CXCL8/10/11, which mediate IFNγ-derived recruitment of CXCR3^+^ T and NK cells; and CCL3, CCL4, CCL8, CXCL8 and CCL20, which recruit monocytes, macrophages, T cells, and NK cells. Evidence of immune checkpoint regulation and exhaustion was also observed, with high abundance of co-inhibitory and metabolic checkpoint molecules (PDCD1/PD-1, PDCD1LG2/PD-L2, IDO1 and KYNU; **Fig. 2B, C, Fig. S9**). In contrast, the 142 proteins that were more abundant but only common between Ethiopia and Sudan formed sparse networks, mostly associated with innate immune sensing and apoptosis (TLR2, TLR4, CASP1/7/8); as well as protein folding and MAPK stress signalling (AHSA1, DNAJA4, MAP2K1, MAPK9, PNPT1) (**Fig. S10**). Gene Ontology, KEGG and Reactome Over Representative Analysis (ORA) supported the main findings observed in the STRING network analysis (**Table S3**).

**Figure 2:**
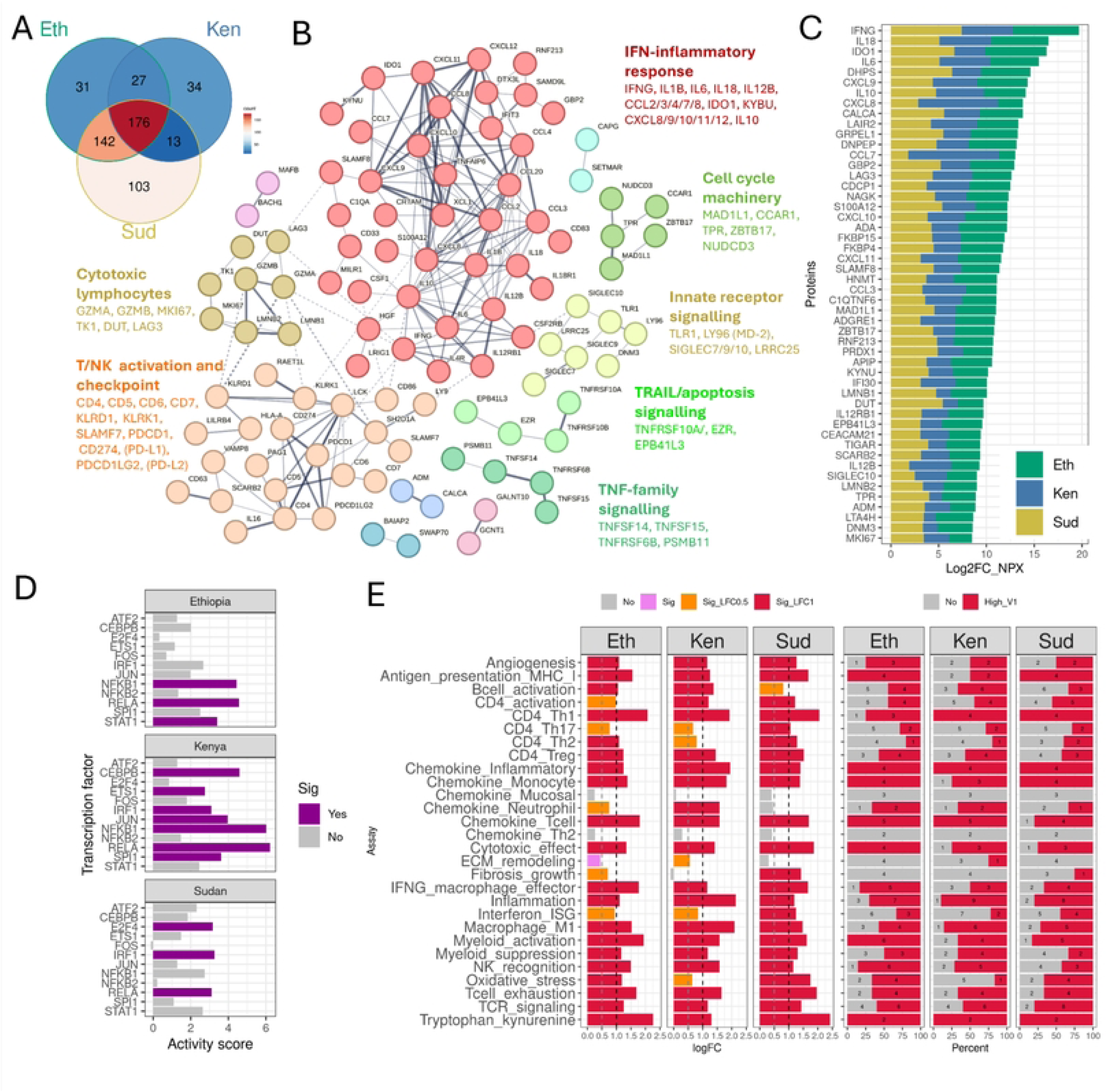
Immune and metabolic hallmarks of VL across East Africa. **A)** Venn diagram showing the intersections of proteins up-regulated in pre-treatment (V1) when compared to HV across the three countries. Left: Ethiopia (Eth), Right: Kenya (Ken) and Bottom: Sudan (Sud). The colour scale from blue to red represents the proportion of proteins in each section of the diagram. **B)** STRING network analysis of the 176 proteins that were upregulated in V1, across the three countries. A larger version of this figure can be seen at <u>Fig. S9</u>. **C)** Top50 proteins upregulated in V1. Proteins are ordered by the combined Log2FC values among the three countries. Each colour represents a different country: Green - Ethiopia, Blue - Kenya, Yellow - Sudan, and the bar width is proportional to the Log2FC values in each country. **D)** Transcription factor activity. Transcription factors that had its activity significantly increased in V1 when compared to HV are in purple. **E)** Module score analysis. Each row represents an individual module and each column represents a country. **Left:** Log2FC of the V1-HV module expression comparison. Bars in gray, pink, yellow and red represent, respectively, not-significant, significant (p-value < 0.05); higher expression (p-value <0.05 and log2FC > 0.5) and top highly expressed (p-value <0.05 and log2FC > 1) modules. The vertical gray and black bars denote, respectively, log2FC values of 0.5 and 1. **Right:** Number of proteins within each module that were significantly overexpressed in V1 compared with HV (p-value <0.05 and log2FC > 1). Red bars represent overexpressed proteins, whereas gray bars represent non-significant proteins. Numbers within the bars indicate the corresponding counts of significant and non-significant proteins.

To broaden the generalizability of this observation, we examined an Olink Reveal dataset derived from Indian VL patients and matched HV, confirming that this Th1/cytotoxic inflammatory response was broadly consistent in VL patients in East Africa and South Asia (**Fig. S11**, **Table S4**).

We next sought to identify upstream regulatory pathways linked to the core protein profile identified in East African patients. Transcription factor activity analysis using the DoRothEA database and univariate linear models (ULM) identified the NF-κB-associated transcription factors NFKB1 as consistently relevant across all three countries; and RELA in Ethiopia and Kenya (**Fig. 2D**, **Fig. S12**, **Tables S5**, **S6**). The transcription factors JUN, ETS1, SPI1 and CEBPB were significantly enriched only in Kenyan patients, suggestive of strong myeloid cell differentiation and inflammatory activation.

To further characterize the variations in immune response, we developed custom modules representing key immune and metabolic processes (see Methods; **Fig. S13**, **Table S7**). Prominent modules highly expressed at V1 included CD4_Th1, Cytotoxic_effect, Tcell_exhaustion and Tryptophan_kynurenine, consistent with the STRING network analysis (**Fig. 2E**). Taken together, these findings indicate that VL patients exhibit a systemic Th1-cytotoxic inflammatory response, recruitment of monocytes/macrophages, neutrophils, T cells and NK cells, and concurrent activation of immunoregulatory and exhaustion-associated pathways.

### Treatment decreases IFNy-driven immune activation, with residual inflammation at EoT

To assess the impact of treatment in these patient cohorts, we compared protein abundances before (V1) and after (V2) treatment, showing a global reduction in the abundance of immune-related proteins following therapy in all three countries, though this was noticeably less pronounced in Kenya (**Fig. 3**, **Fig. S14**). For proteins that decrease in expression with treatment in at least one country, 71/278 (∼25%) were shared between the three countries. Another 112 (∼40%) were only shared between Ethiopia and Sudan, more than were shared between Kenya and Ethiopia (8; ∼3%) or Kenya and Sudan (5; ∼2%) (**Fig. 3A**). The difference in Kenyan patient response was even more pronounced when comparing EOT (V2) and HV, where Kenya-exclusive highly expressed proteins exceeded all country-shared highly expressed proteins (104 vs. 47; **Fig. 3B**, **Fig. S15**). These data suggest that in addition to a core protein signature associated with treatment response and clinical cure, restoration to homeostasis in Kenyan patients was more muted compared to the other countries, as also inferred from our trajectory analysis (**Fig. 1**).

**Figure 3:**
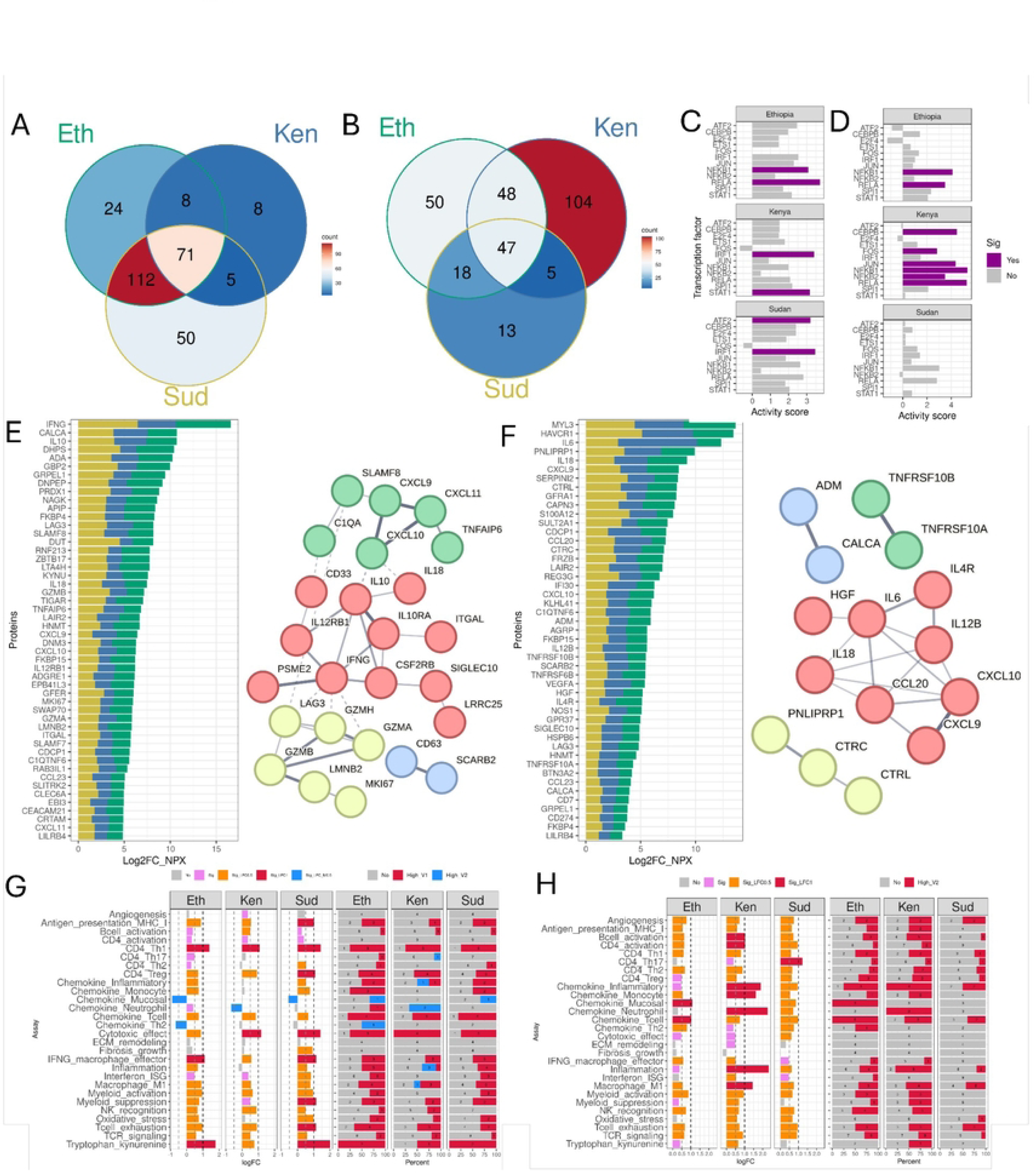
Treatment impact on VL patients across East Africa. **A-B**: Venn diagram showing the intersections of proteins up-regulated in: **A)** pre-treatment (V1) when compared to post-treatment (V2); and **B)** post-treatment (V2) when compared to healthy volunteers (HV); across the three countries **Left**: Ethiopia, **Right**: Kenya and **Bottom**: Sudan. The color scale from blue to red represents the proportion of proteins in each section of the diagram. **C-D**: Transcription factor activity. Transcription factors (TF) that had its activity significantly increased in **C)** V1 when compared to V2; and **D)** V2 when compared to HV. Significant relevant TFs are in purple. **E-F) Left**: STRING network analysis of the proteins that were upregulated across the three countries. **Right** Top50 upregulated proteins. Proteins are ordered by the combined Log2FC values among the three countries. Each colour represents a different country: Green - Ethiopia, Blue - Kenya, Yellow - Sudan, and the bar width is proportional to the Log2FC values in each country. **E)** 71 proteins downregulated in V2 compared to V1; **F)** 47 proteins upregulated in V2 when compared to HVs. A larger version of this figure can be seen at <u>Fig. S16</u>. **G-H)** Module score analysis. Each row represents an individual module and each column represents a country. **G)** V1 versus V2 comparison. **Left**: modules classified as non-significant (gray), V1 significant (pink; p < 0.05), V1-enriched (yellow; p < 0.05 and log₂FC > 0.5), highly V1-enriched (red; p < 0.05 and log₂FC > 1), or V2 significant (blue; p < 0.05). **Right**: number of proteins within each module classified as V1-enriched (red), non-significant (gray), or V2-enriched (blue). **H**) V2 versus HV comparison. Left: modules classified as non-significant (gray), V2 significant (pink; p < 0.05), V2-enriched (yellow; p < 0.05 and log₂FC > 0.5), or highly V2-enriched (red; p < 0.05 and log₂FC > 1). Right: number of proteins within each module classified as V2-enriched (red) or non-significant (gray). Vertical gray and black lines indicate log₂FC thresholds of 0.5 and 1, respectively. Numbers within bars indicate the corresponding protein counts.

The core treatment response in all countries included a partial resolution of IFNγ-associated immune activation, with reduction in the abundance of proinflammatory cytokines (including IFNγ, IL-12RB1, IL-18 and the TNF-inducible hyaluron-binding protein TNFAIP6), CXCR3 chemokine axis cell recruitment (CXCL9, CXLC10, CXCL11) and several proteins involved in cytotoxic lymphocyte activation and function (including GZMA, GZMB and GZMH; **Fig. 3E**, **Fig. S16**, **Table S8**). Parallel reductions in IL-10, IL-10RA, LAG3, and KYNU were also observed, suggesting attenuation of compensatory immunoregulatory and exhaustion-associated pathways consistent with an overall decrease in immune stimulation. While the reduction in core treatment-response was conserved among countries, Ethiopian and Sudanese patients exhibited marked reductions in inflammatory, myeloid and lymphoid-associated proteins and kinases (IL-6, IL-1B, IL-15, CASP1, CASP8 and MAPK9, MAP2K1) and proteins involved in the mitochondrial function (SDHA, SDHB, NFU1 and NDUFA5), which were not observed in Kenyan patients (**Fig. S17**).

Despite the substantial resolution of the Th1/cytotoxic inflammatory response following VL treatment, immune homeostasis was not fully restored at EOT. Abundance of Th1-associated cytokines (IL-12B and IL-18), the inflammatory cytokine IL-6, and chemokines involved in lymphoid and myeloid cell recruitment (CXCL9, CXCL10, and CCL20) in all countries, and the master regulators NFKB and RELA in Ethiopia and Kenya suggests persistent cellular immune activation at EOT (**Fig. 3 D, F**, **Fig. S16**, **Table S9**). Kenya also exhibited persistent monocyte and neutrophil recruitment (CCL7, CXCL1, CXCL6, CXCL8), the pro-inflammatory cytokine IL-1B and proteins associated with lymphocyte recruitment, antigen presentation, and adaptive immune responses (IL-4, CD4, IL-16, and HLA-A) (**Fig. S18**). These changes were accompanied by enrichment of transcription factors associated with inflammatory signalling, cellular stress responses, and myeloid cell activation, including (e.g. CEBPB, NFKB2, FOS and JUN; **Fig. 3 D**). Protein module analysis also supported a reduction in IFNγ-associated immune activation, cytotoxicity and compensatory pathways, and a persistent inflammation with myeloid and neutrophil recruitment in Kenya at EOT, with some country-specific module variations (**Fig. 3G and H**).

Taken together, these results indicate that successful VL treatment results in a partial reduction of the Th1/cytotoxic inflammatory state that characterizes active VL. The more extensive decline in abundance of inflammatory and interferon-responsive proteins in Ethiopia and Sudan compared with Kenya further suggests that regional factors may influence the magnitude or timing of immune restoration after treatment.

### Proteomic differences across East African countries

Directly comparing protein abundance profiles at admission and after treatment reinforced the assertion that patients in Kenya had stronger inflammatory responses at presentation and sustained monocyte and neutrophil recruitment at EOT, when compared to Sudan and Ethiopia (**Fig. 4**, age adjustment results in **Fig. S19**, **Table S10**). To identify proteins that were preferentially more abundant in patients in a given country, we identified proteins that were more abundant relative to both other countries (log2FC > 0.5, adjusted p-value < 0.05) and more abundant than in HV from the same country (log2FC > 1, adjusted p-value < 0.05). Ethiopian patients had few preferentially more abundant proteins (7 at V1, 1 at V2), compared to Kenya (32 and 45, respectively) and Sudan (70 and 13, respectively; **Fig. 4**).

**Figure 4:**
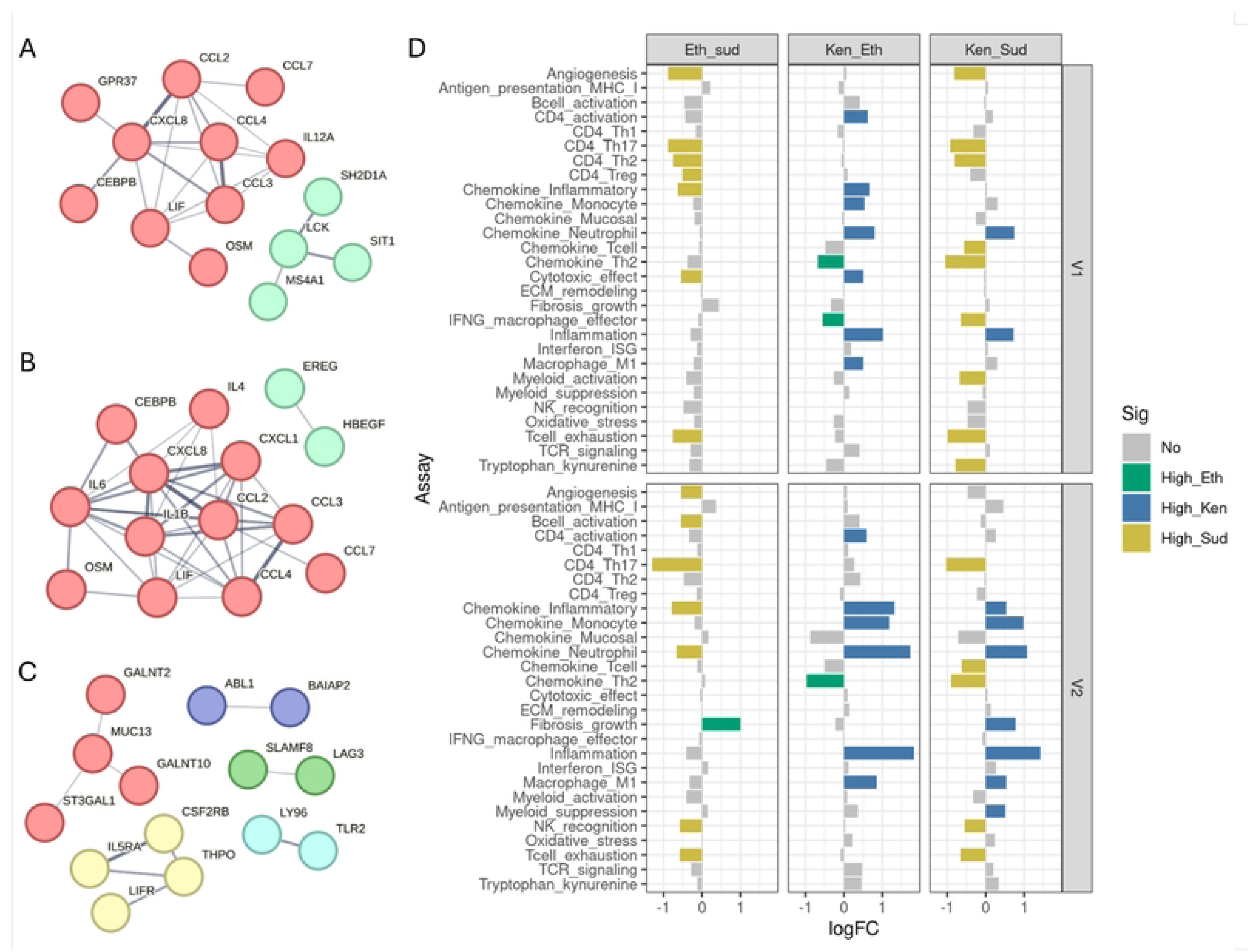
Differences in protein expression among countries at admission and at the end of treatment. **A-C) Left**: STRING db networks from the exclusive proteins that were also higher than the HV for the same country. (**A**) Kenya V1; (**B**) Kenya V2 and (**C**) Sudan V1. No significant STRING networks were observed for Ethiopia (V1 and V2) and Sudan V2. (**D**) Log2FC of the module comparison among countries and among stages. Coloured bars represent modules with significant differences among countries (log2FC > 0.5 and adjusted p-value < 0.05), where high in Ethiopia (High-Eth), high in Kenya (High_Ken) and high in Sudan (High_Sud) are represented respectively in green, blue and gold. Each column corresponds to a country-country comparison, from left to right Ethiopia x Sudan; Kenya x Ethiopia and Kenya x Sudan. The top and lower boxes represent the patient stages, respectively pre-treatment (V1) and post-treatment (V2). The X axis represents the log2FC. Volcano plots can be seen at <u>Fig. S19</u>.

STRING analysis of proteins more abundant in Kenyan patients confirmed a prominent inflammatory myeloid recruitment signature at admission (CXCL8, CCL2, CCL3, CCL4 and CCL7), together with evidence of Th1-polarization (IL-12A) and emergency myelopoiesis and granulopoiesis (CEBPB) [31] (**Fig. 4A**). These features were substantially retained at EOT and accompanied by heightened abundance of acute and inflammatory mediators (IL-1B, IL-6 and OSM) (**Fig. 4B**). Module analysis broadly confirmed these observations (**Fig. 4D**, **Fig. 5A-D**, **Fig. S20**). Of note, chemokines such as CCL20, CCL23, CCL8 and CX3CL1, which recruit broader leukocyte populations and are often associated with tissue immune surveillance or chronic inflammatory responses, were not similarly enriched in Kenyan patients. In Sudanese patients, the STRING analysis of differentially expressed genes only captured a few sparse clusters in V1, which is probably due to the lower sample size for this set (**Fig. 4C**). Module analysis supported a more prominent Th17 response and exhaustion in Sudan (**Fig. 5E, F**).

**Figure 5:**
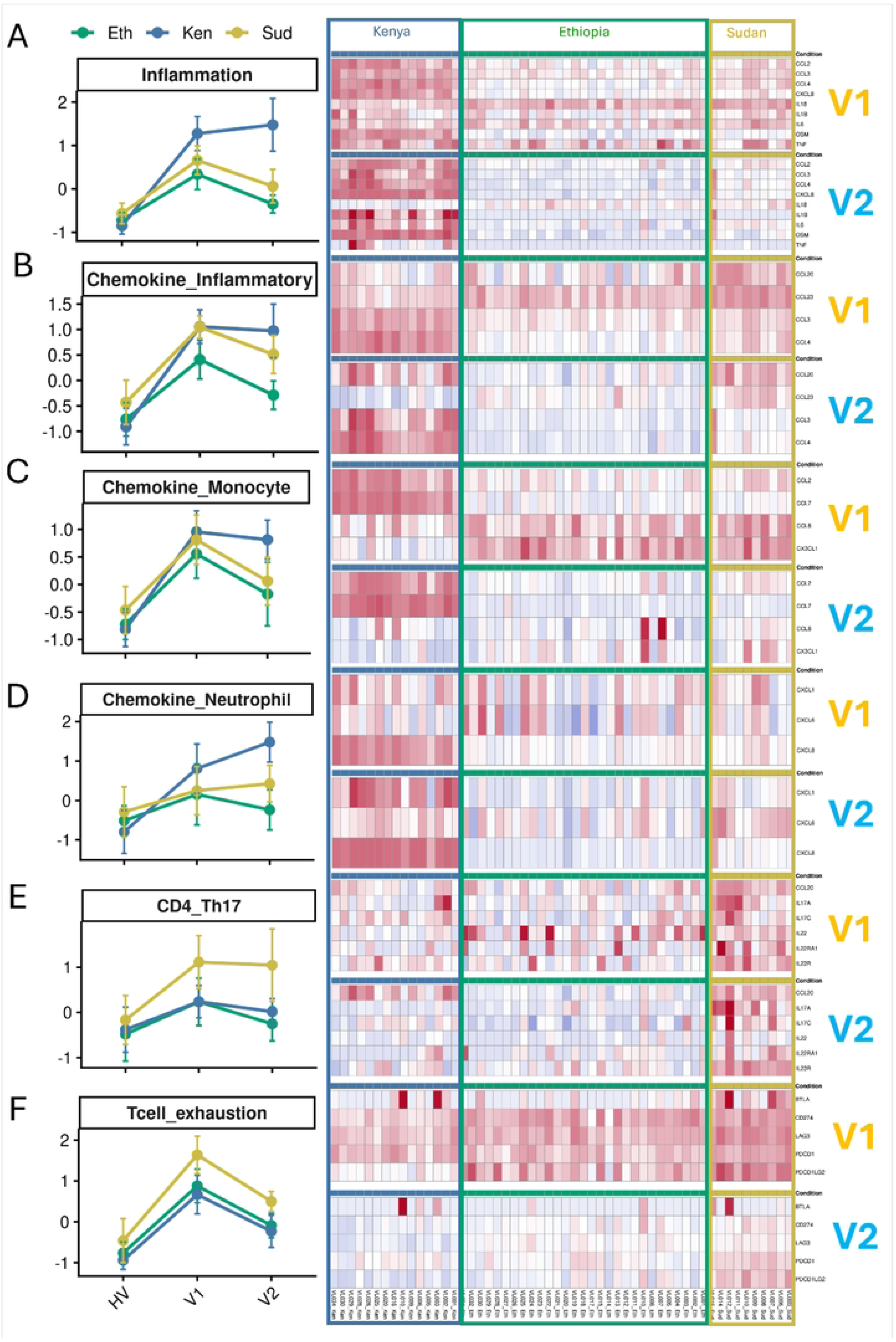
Main inter-country variable modules change across disease stages and patients. **Left**: Lineplot showing the mean (dot) and standard deviation (whiskers) of the Z-scored scaled NPX values from all patients from each country’s healthy volunteers (HV), pre-treatment (V1) and post-treatment (V2) groups. Ethiopia, Kenya and Sudan are respectively represented in green, blue and gold. **Right**: Heatmap showing the individual value for each patient, for each marker within each module. Each row represents a protein, while each column represents a patient. The colourstrip and boxes highlight the sample country of origin. For each panel, the top and bottom heatmaps represent, respectively, pre-treatment (V1) and post-treatment samples (V2). **A)** Inflammation; **B)** Chemokine inflammatory; **C)** Chemokine_Monocyte; **D)** Chemokine_Neutrophil; **E)** CD4_Th17 and **F)** Tcell_exhaustion. Tendency plots for all modules can be seen at <u>Fig. S20</u>.

Taken together, these results highlight distinct country-specific immune signatures in VL. Kenyan patients exhibited a sustained inflammatory and myeloid recruitment profile, whereas Sudanese patients showed signatures associated with lymphocyte activation and immune regulation. These differences suggest regional variation in the host immune response to VL.

### Associations between protein levels and patient clinical traits

Finally, we used limma linear modelling with empirical Bayes moderation accounting for within-subject correlation, patient age and country of origin to evaluate potential associations between proteomic variations and patient clinical traits, with focus on Albumin and ALT levels, two of the main markers assessed in VL patients to evaluate cure (**Fig. 6, Table S11**). Results for other clinical traits can be seen in **Fig. S21**. We used samples where both pre-treatment and post-treatment clinical data from the same patient was available (28, 8 and 10 samples from Ethiopia, Kenya and Sudan, respectively). Given the small sample size and uneven distribution, these analyses should be viewed as exploratory.

**Figure 6:**
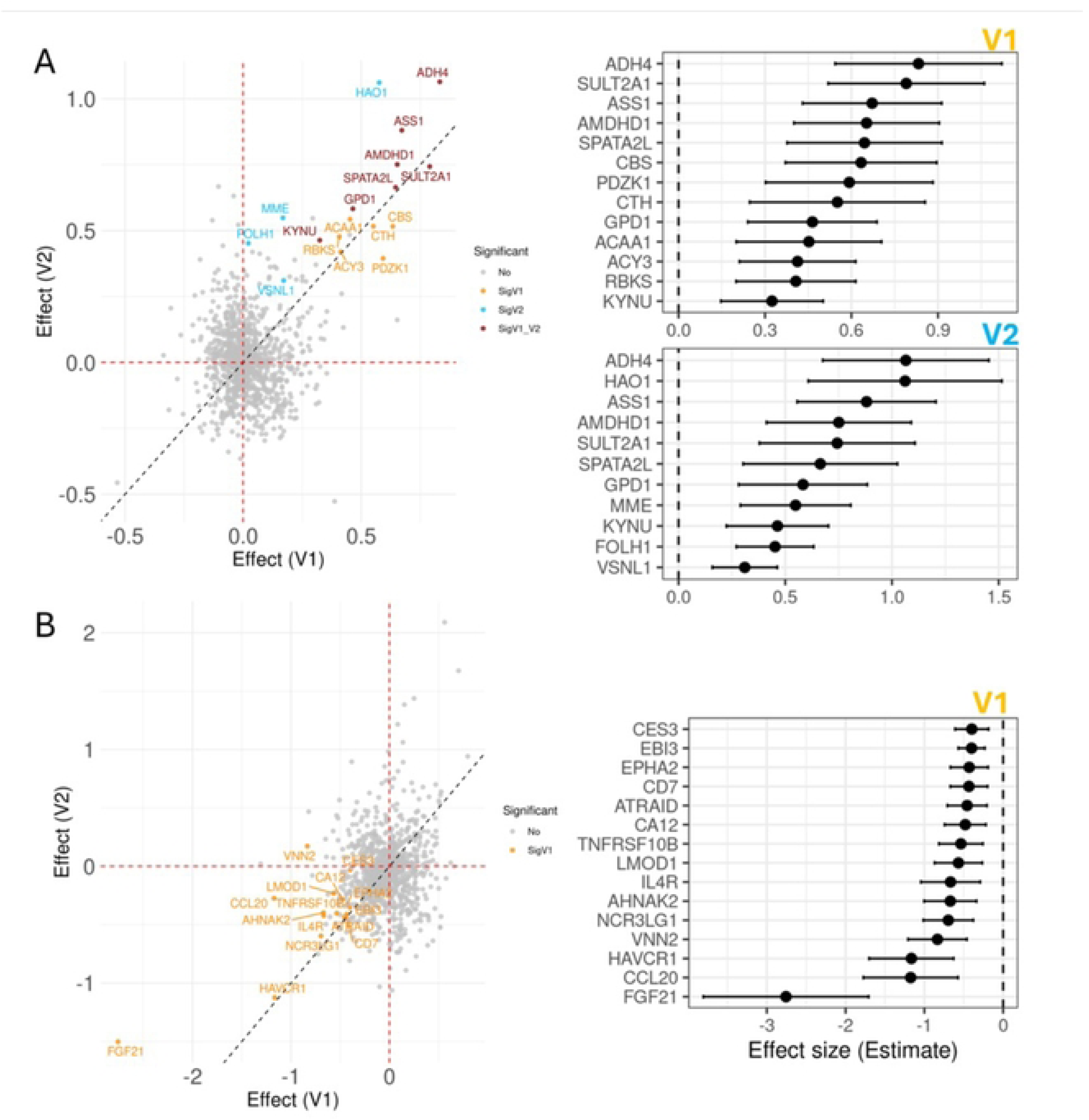
Associations between clinical traits and Olink protein levels in VL patients before and after treatment. **Left**: Dotplot representing the effect association between a trait and all proteins in the Olink panel. Each dot corresponds to a protein, where proteins significantly associated with a given trait (adjusted p-value < 0.05 and confidence interval not crossing 0) are coloured and named. The colours orange, blue and brown represent, respectively, proteins that were associated with the clinical phenotype only on V1, V2 or in both V1 and V2. The X and Y axis represents, respectively, the association between the protein and the trait in V1 and V2. The dashed red line marks “0” for both axes, where negative or positive values indicate, respectively, inverse and direct correlations. **Right** panel: 95% confidence interval for the association effective size.

We observed 6, 4 and 7 proteins significantly associated with high ALT levels in respectively V1, V2 and simultaneously in V1 and V2 (**Fig. 6A**). Higher ALT levels in VL patients are associated predominantly with the circulant levels of liver-enriched enzymes: ADH4 on both stages and HAO1 only on V2, which might represent evidence for liver damage or dysregulation. ASS1, Argininosuccinate Synthase 1, an enzyme involved in arginine biosynthesis was also associated with higher ALT levels on both stages. This enzyme is important for resynthesizing arginine, ensuring sufficient substrate supply for NO production for prolonged mycobacteria infection [32, 33]. We also observed association with KYNU, a component of the tryptophan–kynurenine pathway that can lead to T cell suppression and parasite survival [34, 35]. Other potential associations were related to amino acid metabolism (ACY3, CBS, CTH), lipid and energy metabolism (ACAA1, GPD1), steroid and bile acid metabolism (SULT2A1).

We observed a total of 15 proteins associated with lower Albumin levels (**Fig. 6B**). The highest effect association was with Fibroblast growth factor 21 (FGF21), which is mainly produced by the liver and adipose tissue, and was proposed as a marker for acute liver injury [36–38]. There was also association with HAVCR1 (KIM-1, TIM-1), a marker of systemic illness and kidney injury [39]. Some immune related proteins were also associated with lower albumin levels, such as CCL20 that recruits immature dendritic cells, Th17 and B cells; IL-4R, which supports Th2 cell differentiation; and TNFRSF10B, a TNF receptor that transduces apoptosis signals.

## Discussion

The control of VL in East Africa remains a significant challenge, despite the advent of new drugs and drug combinations with promising clinical efficacy. In this context, understanding the pathophysiology of disease and treatment response at a systems level becomes increasingly important for capturing disease complexity and the nature of drug-immune-metabolism interactions. Whilst transcriptomic studies in VL patients before and after treatment have been widely reported, the identification of proteomic changes has fallen behind. The analysis reported here provides the first comprehensive targeted evaluation of the plasma proteome before and after treatment of VL patients across multiple sites in East Africa.

Plasma proteomics studies are being increasingly applied to understand the pathophysiology of human disease, with a growing number of examples from the field of infectious disease. Recent studies have revealed new features associated with NK cell activity in malaria [40] and HIV [41], reported the effects of BCG vaccination on systemic inflammation in neonates [42], identified persistent inflammatory markers in HIV-1 patients on anti-retroviral therapy [43] and identified divergent profiles in infants infected with SARS-CoV-2 and RSV [44]. However, the value of plasma proteomics for evaluating treatment response has been largely confined to studies in cancer, autoimmune or allergic disease patients responding to biological therapies [45–47], or patients with viral infection. For example, in people living with HIV, proteomic aging was shown to be accelerated and linked to metabolic complications [48] and baseline metabolic profiles have been associated with treatment response in people living with HIV [49, 50].

The most comprehensive study to date addressing proteomic changes during parasitic infection and in response to treatment was provided by Lautenbach et al, reporting on the response in a cohort of 72 adults travellers returning to Sweden, with diagnosed symptomatic *P. falciparum* infection [22]. In this study, 703 differentially abundant proteins were identified comparing acute disease and samples collected 10 days after initiation of treatment (parasitological cure) and at 12 months post treatment (regarded as baseline or “healthy”). Many of these proteins were immune related and linked to known features of malaria pathogenesis, notably interferon responses and cytotoxicity. Whilst most differentially abundant proteins rapidly declined to baseline on treatment, 43 remained elevated including IL-10, killer cell immunoglobulin-like receptors (KIR3DL1, KIR2DL3) and CD22. Erythropoietic (EPO) increased post treatment, suggestive of immune restoration. This study has parallels to our current work in VL patients, but with some important differences. First, our cohorts were recruited and treated in four endemic countries in East Africa, allowing stratification of response by country. Although emphasis is placed during our analysis on the matched paired comparison of patients pre and post treatment, we included convenience controls to provide a surrogate baseline. Whilst not fully matched demographically to the patient groups, this approach avoided potential confounders (e.g. coinfection) associated with long term follow up in an endemic setting. Second, whilst standard of care (17 days sodium stibogluconate and paromomycin) led to clinical cure in all patients, it is recognised that this is unlikely to be sterile cure and a low grade sub-clinical parasite burden may persist, driving persistent inflammation. Nevertheless, our data show many similarities with those reported in severe malaria, likely attributable to common pathophysiological pathways and clinical symptoms. Interferon-, TNF- and cytotoxicity-related pathways were similarly impacted and declined post treatment. Of note, these findings were consistent in an Olink dataset derived from a small cohort of Indian VL patients, suggesting this is a generalised feature of VL. Evidence of liver damage was observed, including increased abundance of HAO1, as seen in malaria [22].

A major yet unexpected finding from our study was the difference in proteomic response to treatment observed in patients from different countries in East Africa. At the level of the measured proteome, the diverse response of Kenyan patients compared to that of patients from Ethiopia and Sudan was striking when visualised by trajectory analysis. This clearly demonstrated both diversity of differentially abundant proteins at presentation and more importantly a difference in the rate of recovery towards homeostasis. Despite a core signature of 176 proteins identified as being associated with VL in all countries, additional differentially abundant proteins were observed as a country level that shaped this trajectory. Amongst these, prominent and sustained myeloid cell-focused inflammatory response pathways were observed in Kenyan patients. This included multiple myeloid cell-derived chemokines (e.g. CCL2, CCL3, CCL4, CXCL8) as well as multiple chemokines that drive myeloid cell recruitment (incl. CCL3, CCL4). Transcription factor analysis indicated that myeloid-selective transcription factors (SPI1 and CEBPB) were also most prominent in Kenyan patients. Although the response in Kenyan patients was most striking, other country specific differences in immune modules were also observed that did not appear to affect overall trajectory of the treatment response. For example, pathways associated with CD4^+^ Th17 cells and with exhausted T cells (including LAG3, PD1/PD-L1) were more prominent in Sudanese patients despite there being no major difference in response trajectory between Sudan and Ethiopia. Nevertheless, and despite these differences in protein abundances, patients across these cohorts/countries responded similarly to treatment, relapses were few and the incidence of PKDL in the Sudanese cohort was low. Although age was a clear variable across cohorts, these differences all survived age adjusted analysis. Although we cannot formally explain the reason for these country-specific differences in response trajectory, they likely arise from confounders that were not included in our current analysis (e.g. prior infection history, diet, host and / or parasite genetics). Although clinical trials of new anti-leishmanial drugs have shown country-specific variations in efficacy [51, 52], it is reasonable to conclude that the immune response differences noted in the cohorts studied here had limited impact on clinical presentation or response to treatment with SSG/PM. It remains to be determined whether these or other immune response differences may impact treatment outcome following the introduction of new drugs / drug combinations or whether their effects might have been observed in longitudinal follow up studies monitoring more diverse aspects of health.

Proteomic analysis has additional value in potentially identifying biomarkers for differential diagnosis and / or stratification of diseases. Amongst the 11 proteins (TNFRSF1B, LAYN, IL-2RA, B4GALT1, SPINK1, CALCA, TNFRSF10B, ICAM1, IFNGR1, HMOX1) identified by Lautenbach et al as enriched in severe malaria compared to other febrile conditions of infectious origin (*E. coli*, *Salmonella*, *Shigella*, *Mycoplasma pneumoniae*, Dengue virus, *Campylobacter*, Influenza A/ B), four proteins are present in the Olink Reveal panel used here (CALCA, IFNGR1, LAYN, TNFRSF10B). We found two (CALCA and TNFRSF10B) were consistently observed at presentation in VL patients across all three EA countries, where CALCA was also elevated in patients from India. CALCA is a precursor protein that is alternatively spliced and processed to generate procalcitonin (PCT) and calcitonin-gene-related peptide (CGRP). Elevated PCT levels have been observed in VL patients in Iran, independently of secondary bacterial infection [53]. CGRP is a sensory neuropeptide that has been reported to have an immune regulatory role and affect nociception in murine models of cutaneous leishmaniasis [54, 55] but its role if any in VL is yet to be established. TNFRSF10B also known as Death Receptor-5 (DR5) or TRAIL-R2 is a type I transmembrane protein of the TNF receptor superfamily, acts as a central hub in the regulation of apoptosis by binding its ligand TRAIL [56], and is a target for biologic therapy in cancer [57]. DR5 has been linked to CD4^+^ T cell death and immunosuppression in murine models of VL [58]. There was also evidence of higher protein abundances for LAYN and IFNGR1 in VL patients, albeit not similarly strong in all countries. LAYN was significantly elevated at presentation in Ethiopia and Kenya and in Sudan at a lower cutoff (log2FC = 0.89); and in Indian patients (log2FC=0.67). IFNGR1 was elevated in Ethiopia at admission, and also in Kenya (at Log2FC = 0.4), but not in Sudan or India. IFNGR1 is the Interferon gamma receptor 1 subunit, which when combined with IFNGR2 becomes the main receptor for IFNγ. IFNγ signalling promotes Th1-type immune responses, and macrophage activation, and polymorphisms in the receptor have been associated with PKDL susceptibility in Sudan [59]. These data further highlight the potential for country-specific differences in host response to VL.

The relatively small sample size of this study and the limited availability of quantitative clinical data made it challenging to find significant associations between proteomic changes and other clinical traits. In our exploratory analysis, we were able to identify a small number of proteins that were associated with elevated plasma ALT and lower plasma albumin (15 proteins). Whilst this included a number of proteins with biologically plausible relationships to tissue damage and fibrosis and which therefore might serve as additional biomarkers of diseases, their identification needs to be treated with caution and confirmed in more extensive studies.

The study has some additional limitations. Some samples failed QC and were excluded from analysis, generating country cohorts of unequal size. Limited blood / serum was available for follow on analysis or for additional phenotyping / scRNA-seq of immune cells. All patients receiving treatment were cured at day 17 and the number of relapses was very low [1], negating the identification of biomarkers associated with treatment outcome.

In summary, our data illustrate both the complexity of the immune and metabolic changes associated with VL and the potential for selective country-specific differences in response. Whilst these changes do not appear to affect clinical outcome on treatment, they nevertheless point to subtle differences in VL immune responses that warrant further investigation. The data provided here will also provide a valuable resource for future comparative studies across more diverse different geographic regions, for linkage studies that assess the impact of parasite genotype on immune / metabolic response and for use during the evaluation of new therapies against VL.

## Acknowledgments

We thank all patients and their families for agreeing to participate and their collaboration. The authors thank all members of the field teams at the study sites of Chemolingot, Amudat, Doka and Gondar, including clinicians, nurses, laboratory technicians, and the local communities for their contributions to the study. We also acknowledge the Ministries of Health of Kenya and Sudan; the Ministry of Education in Ethiopia and the National Council for Science and Technology in Uganda for their support. The authors thank the African Centre for Community Investment in Health (ACCIH), a partner to the Chemolingot Sub County Hospital, Kenya, and the County Government of Baringo for allowing them to undertake the study at the Chemolingot hospital. Disclaimer. This manuscript is published with permission from the Director of Kenya Medical Research Institute. The authors also thank the members of the Kala-Azar Medical Research Center (KAMRC) in Muzaffarpur, Bihar, India. We are grateful to members of the Scientific Advisory Board, especially Prof N.K. Ganguly, for the encouragement to pursue this proteomic analysis.

## Data availability

The R script used for data analysis can be obtained from GitHub: https://github.com/jaumlrc/Immstat_cure_OlinkProteomics.

## Author contributions

**Conceptualization**: Peter O’Toole, Charles J.N. Lacey, Jane Mbui, Asrat M. Hailu, Paul M. Kaye, Margaret Mbuchi, Ahmed M Musa, Joseph Olobo, Christian R. Engwerda.

**Data curation**: Ayenew Addisu, Alice Bayiyana, Joao Cunha, Daniel Matano, Brima M. Younis, Karen Hogg.

**Formal analysis**: Joao Cunha, Rebecca Wiggins.

**Funding acquisition**: Peter O’Toole, Charles J.N. Lacey, Jane Mbui, Asrat M. Hailu, Paul M. Kaye, Margaret Mbuchi, Ahmed M Musa, Joseph Olobo.

**Investigation**: Ayenew Addisu, Alice Bayiyana, Joao Cunha, Daniel Matano, Brima M. Younis, Karen Hogg, Rebecca Wiggins, Wilson Biwott, Finnley Osuna, Christine Ichugu, Ayalew Jejaw Zeleke, Eleni Ayele, Obondo James Sande, Eltahir A.G. Khalil, Hussam M.H. Ibrahim, Mahmoud A. Mahmoud, Ahmed I.B. Zakaria, Brenda Adiko, Peter O’Toole, Charles J.N. Lacey, Jane Mbui, Asrat M. Hailu, Paul M. Kaye, Margaret Mbuchi, Ahmed M Musa, Joseph Olobo, Christian R. Engwerda, Shyam Sundar, Jessica Engel, Rahul Tiwari, Rajiv Kumar.

**Methodology**: Joao Cunha, Karen Hogg, Peter O’Toole, Charles J.N. Lacey, Jane Mbui, Asrat M. Hailu, Paul M. Kaye, Margaret Mbuchi, Ahmed M Musa, Joseph Olobo, Christian R. Engwerda.

**Project administration**: Rebecca Wiggins, Flavia D’Alessio.

**Resources**: Peter O’Toole, Charles J.N. Lacey, Jane Mbui, Asrat M. Hailu, Paul M. Kaye, Margaret Mbuchi, Ahmed M. Musa, Joseph Olobo.

**Supervision**: Joao Cunha, Karen Hogg, Peter O’Toole, Charles J.N. Lacey, Jane Mbui, Asrat M. Hailu, Paul M. Kaye, Margaret Mbuchi, Ahmed M Musa, Christian R. Engwerda.

**Validation**: Rebecca Wiggins, Paul M. Kaye.

**Visualization**: Joao Cunha.

**Writing – original draft**: Joao Cunha.

**Writing – review & editing**: Ayenew Addisu, Alice Bayiyana, Joao Cunha, Daniel Matano, Brima M. Younis, Karen Hogg, Rebecca Wiggins, Wilson Biwott, Finnley Osuna, Christine Ichugu, Ayalew Jejaw Zeleke, Eleni Ayele, Obondo James Sande, Eltahir A.G. Khalil, Hussam M.H. Ibrahim, Mahmoud A. Mahmoud, Ahmed I.B. Zakaria, Brenda Adiko, Peter O’Toole, Charles J.N. Lacey, Jane Mbui, Asrat M. Hailu, Paul M. Kaye, Margaret Mbuchi, Ahmed M Musa, Joseph Olobo, Christian R. Engwerda, Rajiv Kumar, Rahul Tiwari, Jessica Engel, Shyam Sundar

## References

1. Global leishmaniasis surveillance updates 2023: 3 years of the NTD road map. 2024.

2. Burza S, Croft SL, Boelaert M. Leishmaniasis. Lancet (London, England). 2018;392. doi:10.1016/S0140-6736(18)31204-2

3. Nagi N. Bangladesh eliminates visceral leishmaniasis. Lancet Microbe. 2024;5(5):e420. doi: 10.1016/S2666-5247(24)00028-4.

4. Pandey DK, Alvar J, den Boer M, Jain S, Gill N, Argaw D, et al. Kala-azar elimination in India: reflections on success and sustainability. Int Health. 2025;17(4):416–22. doi: 10.1093/inthealth/ihaf013. PubMed Central PMCID: PMCPMC12212201.

5. Strategic framework elimination visceral leishmaniasis public health problem eastern Africa2023 2023.

6. Alvar J, Vélez ID, Bern C, Herrero M, Desjeux P, Cano J, et al. Leishmaniasis worldwide and global estimates of its incidence. PLoS One. 2012;7(5):e35671. doi: 10.1371/journal.pone.0035671. PubMed Central PMCID: PMCPMC3365071.

7. Ashwin H, Seifert K, Forrester S, Brown N, MacDonald S, James S, et al. Tissue and host species-specific transcriptional changes in models of experimental visceral leishmaniasis. Wellcome Open Res. 2018;3:135. doi: 10.12688/wellcomeopenres.14867.1. PubMed Central PMCID: PMCPMC6248268.

8. Forrester S, Goundry A, Dias BT, Leal-Calvo T, Moraes MO, Kaye PM, et al. Tissue specific dual RNA-seq defines host-parasite interplay in Murine visceral leishmaniasis caused by Leishmania donovani and Leishmania infantum. Microbiol Spectr. 2022;10(2):e0067922. doi: 10.1128/spectrum.00679-22. PubMed Central PMCID: PMCPMC9045295.

9. Maruyama SR, Fuzo CA, Oliveira AER, Rogerio LA, Takamiya NT, Pessenda G, et al. Insight into the long noncoding RNA and mRNA coexpression profile in the human blood transcriptome upon Leishmania infantum infection. Front Immunol. 2022;13:784463. doi: 10.3389/fimmu.2022.784463. PubMed Central PMCID: PMCPMC8965071.

10. Fakiola M, Singh OP, Syn G, Singh T, Singh B, Chakravarty J, et al. Transcriptional blood signatures for active and amphotericin B treated visceral leishmaniasis in India. PLoS Negl Trop Dis. 2019;13(8):e0007673. doi: 10.1371/journal.pntd.0007673. PubMed Central PMCID: PMCPMC6713396.

11. Adriaensen W, Cuypers B, Cordero CF, Mengasha B, Blesson S, Cnops L, et al. Host transcriptomic signature as alternative test-of-cure in visceral leishmaniasis patients co-infected with HIV. EBioMedicine. 2020;55(102748):102748. doi: 10.1016/j.ebiom.2020.102748. PubMed Central PMCID: PMCPMC7195535.

12. Caldas A, Favali C, Aquino D, Vinhas V, van Weyenbergh J, Brodskyn C, et al. Balance of IL-10 and interferon-gamma plasma levels in human visceral leishmaniasis: implications in the pathogenesis. BMC Infect Dis. 2005;5(1):113. doi: 10.1186/1471-2334-5-113. PubMed Central PMCID: PMCPMC1343567.

13. Tadesse D, Abdissa A, Mekonnen M, Belay T, Hailu A. Antibody and cytokine levels in visceral leishmaniasis patients with varied parasitemia before, during, and after treatment in patients admitted to Arba Minch General Hospital, southern Ethiopia. PLoS Negl Trop Dis. 2021;15(8):e0009632. doi: 10.1371/journal.pntd.0009632. PubMed Central PMCID: PMCPMC8370634.

14. van den Bogaart E, Talha A-BA, Straetemans M, Mens PF, Adams ER, Grobusch MP, et al. Cytokine profiles amongst Sudanese patients with visceral leishmaniasis and malaria co-infections. BMC Immunol. 2014;15(1):16. doi: 10.1186/1471-2172-15-16. PubMed Central PMCID: PMCPMC4024313.

15. van Dijk N, Carter J, Kiptanui D, Pinelli E, Schallig H. Cytokine profiles, blood parasite load and clinical features of visceral leishmaniasis in West Pokot County, Kenya. Parasitology. 2024;151(7):753–61. doi: 10.1017/S0031182024000751. PubMed Central PMCID: PMCPMC11474019.

16. Torres A, Montero-Calle A, Lozano-Rendal M, Sánchez C, Bernardo L, Solana JC, et al. Proteomics of plasma-derived extracellular vesicles from human patients identifies biomarkers for monitoring visceral leishmaniasis therapy. Front Immunol. 2025;16:1646335. doi: 10.3389/fimmu.2025.1646335. PubMed Central PMCID: PMCPMC12464055.

17. Bag AK, Saha S, Sundar S, Saha B, Chakrabarti A, Mandal C. Comparative proteomics and glycoproteomics of plasma proteins in Indian visceral leishmaniasis. Proteome Sci. 2014;12(1):48. doi: 10.1186/s12953-014-0048-z. PubMed Central PMCID: PMCPMC4179796.

18. Rukmangadachar LA, Kataria J, Hariprasad G, Samantaray JC, Srinivasan A. Two-dimensional difference gel electrophoresis (DIGE) analysis of sera from visceral leishmaniasis patients. Clin Proteomics. 2011;8(1):4. doi: 10.1186/1559-0275-8-4. PubMed Central PMCID: PMCPMC3167202.

19. Greenwood C, Ruff D, Kirvell S, Johnson G, Dhillon HS, Bustin SA. Proximity assays for sensitive quantification of proteins. Biomol Detect Quantif. 2015;4:10–6. doi: 10.1016/j.bdq.2015.04.002. PubMed Central PMCID: PMCPMC4822221.

20. Kirsher DY, Chand S, Phong A, Nguyen B, Szoke BG, Ahadi S. Current landscape of plasma proteomics from technical innovations to biological insights and biomarker discovery. Commun Chem. 2025;8(1):279. doi: 10.1038/s42004-025-01665-1. PubMed Central PMCID: PMCPMC12462477.

21. van der Burgt YEM, de Meijer E, Palmblad M. Brief evaluation of Olink Reveal proximity extension assay for high-throughput proteomics: A case study using NIST SRM 1950 and two spike-in protein standards. J Proteome Res. 2025;24(10):5292–6. doi: 10.1021/acs.jproteome.5c00571. PubMed Central PMCID: PMCPMC12501989.

22. Lautenbach MJ, Wyss K, Yman V, Foroogh F, Satarvandi D, Mousavian Z, et al. Systems analysis of clinical malaria reveals proteomic perturbation and innate-adaptive crosstalk linked to disease severity. Immunity. 2025;58(8):2120–36 e5. Epub 20250714. doi: 10.1016/j.immuni.2025.06.014. PubMed PMID: 40664217.

23. Addisu A, Bayiyana A, Reis Cunha JL, Matano D, Younis BM, Hogg K, et al. Systemic inflammatory markers of visceral leishmaniasis treatment response in East Africa. PLoS Negl Trop Dis. 2026;20(2):e0013749. doi: 10.1371/journal.pntd.0013749. PubMed Central PMCID: PMCPMC12965683.

24. Burns JM, Jr., Shreffler WG, Benson DR, Ghalib HW, Badaro R, Reed SG. Molecular characterization of a kinesin-related antigen of Leishmania chagasi that detects specific antibody in African and American visceral leishmaniasis. Proc Natl Acad Sci U S A. 1993;90(2):775–9. doi: 10.1073/pnas.90.2.775. PubMed Central PMCID: PMCPMC45748.

25. Konopka T. Uniform Manifold Approximation and Projection [R package umap version 0.2.10.0]. 2023.

26. Ritchie ME, Phipson B, Wu D, Hu Y, Law CW, Shi W, et al. limma powers differential expression analyses for RNA-sequencing and microarray studies. Nucleic Acids Res. 2015;43(7):e47. doi: 10.1093/nar/gkv007. PubMed Central PMCID: PMCPMC4402510.

27. Yu G, Wang L-G, Han Y, He Q-Y. clusterProfiler: an R package for comparing biological themes among gene clusters. OMICS. 2012;16(5):284–7. doi: 10.1089/omi.2011.0118. PubMed Central PMCID: PMCPMC3339379.

28. Badia-I-Mompel P, Vélez Santiago J, Braunger J, Geiss C, Dimitrov D, Müller-Dott S, et al. decoupleR: ensemble of computational methods to infer biological activities from omics data. Bioinform Adv. 2022;2(1):vbac016. doi: 10.1093/bioadv/vbac016. PubMed Central PMCID: PMCPMC9710656.

29. Garcia-Alonso L, Holland CH, Ibrahim MM, Turei D, Saez-Rodriguez J. Benchmark and integration of resources for the estimation of human transcription factor activities. Genome Res. 2019;29(8):1363–75. doi: 10.1101/gr.240663.118. PubMed Central PMCID: PMCPMC6673718.

30. Forrester S, Siefert K, Ashwin H, Brown N, Zelmar A, James S, et al. Tissue-specific transcriptomic changes associated with AmBisome® treatment of BALB/c mice with experimental visceral leishmaniasis. Wellcome Open Res. 2019;4:198. doi: 10.12688/wellcomeopenres.15606.1. PubMed Central PMCID: PMCPMC6961418.

31. Hirai H, Zhang P, Dayaram T, Hetherington CJ, Mizuno S-I, Imanishi J, et al. C/EBPbeta is required for ’emergency’ granulopoiesis. Nat Immunol. 2006;7(7):732–9. doi: 10.1038/ni1354.

32. Qualls JE, Subramanian C, Rafi W, Smith AM, Balouzian L, DeFreitas AA, et al. Sustained generation of nitric oxide and control of mycobacterial infection requires argininosuccinate synthase 1. Cell Host Microbe. 2012;12(3):313–23. doi: 10.1016/j.chom.2012.07.012. PubMed Central PMCID: PMCPMC3444824.

33. Rath M, Müller I, Kropf P, Closs EI, Munder M. Metabolism via arginase or nitric oxide synthase: Two competing arginine pathways in macrophages. Front Immunol. 2014;5:532. doi: 10.3389/fimmu.2014.00532. PubMed Central PMCID: PMCPMC4209874.

34. Dey NS, Senaratne S, Somaratne V, Madarasinghe NP, Seneviratne B, Forrester S, et al. Early reduction in PD-L1 expression predicts faster treatment response in human cutaneous leishmaniasis. J Clin Invest. 2021;131(22):e142765. doi: 10.1172/JCI142765. PubMed Central PMCID: PMCPMC8592550.

35. Kim M, Tomek P. Tryptophan: A rheostat of cancer immune escape mediated by immunosuppressive enzymes IDO1 and TDO. Front Immunol. 2021;12:636081. doi: 10.3389/fimmu.2021.636081. PubMed Central PMCID: PMCPMC7940516.

36. Falamarzi K, Malekpour M, Tafti MF, Azarpira N, Behboodi M, Zarei M. The role of FGF21 and its analogs on liver associated diseases. Front Med (Lausanne). 2022;9:967375. doi: 10.3389/fmed.2022.967375. PubMed Central PMCID: PMCPMC9705724.

37. Wu L, Pan Q, Wu G, Qian L, Zhang J, Zhang L, et al. Diverse changes of circulating fibroblast growth factor 21 levels in hepatitis B virus-related diseases. Sci Rep. 2017;7(1):16482. doi: 10.1038/s41598-017-16312-6. PubMed Central PMCID: PMCPMC5705770.

38. Zhang Z, Yuan L, Zhang J, Gu Q, Yan F. The predictive role of FGF21 in acute liver injury caused by bacterial infectious diseases in critical care: A retrospective cohort study. J Inflamm Res. 2025;18:6795–806. doi: 10.2147/JIR.S521327. PubMed Central PMCID: PMCPMC12124309.

39. Song J, Yu J, Prayogo GW, Cao W, Wu Y, Jia Z, et al. Understanding kidney injury molecule 1: a novel immune factor in kidney pathophysiology. Am J Transl Res. 2019;11(3):1219–29. PubMed Central PMCID: PMCPMC6456506.

40. Xi P, Sandoz PA, Lautenbach MJ, Bilev E, Onfelt B, Farnert A, et al. NK cell cytotoxicity is transiently enhanced during acute malaria and modulated by the host microenvironment. JCI Insight. 2026;11(11). Epub 20260421. doi: 10.1172/jci.insight.198687. PubMed PMID: 42013130; PubMed Central PMCID: PMCPMC13313507.

41. Cao WJ, Yang BP, Chen SY, Zhen C, Li XM, Cao XY, et al. Single-cell sequencing reveals NK cell dysregulation during chronic HIV infection and identifies transitional NK cell cytotoxicity related to HIV reservoir decay. EBioMedicine. 2026;130:106381. Epub 20260720. doi: 10.1016/j.ebiom.2026.106381. PubMed PMID: 42475880; PubMed Central PMCID: PMCPMC13393751.

42. Koeken VACM, Nissen TN, Birk NM, Boahen CK, van Crevel R, Kumar V, et al. The effect of BCG vaccination on systemic inflammation in neonates. Vaccine. 2026;75:128241. doi: 10.1016/j.vaccine.2026.128241.

43. Shi Y, Ding C, Zhang Z, Zhang T, Liu J, Huang L, et al. Persistent inflammatory markers existing with HIV-1 reservoirs in antiretroviral therapy treated HIV-1 individuals. Cytokine. 2026;200:157122. doi: 10.1016/j.cyto.2026.157122.

44. Thibodeau A, Mejias A, Nehar-Belaid D, Marches R, Xu Z, Eryilmaz G, et al. A systems immunology approach reveals divergent immune profiles of RSV and SARS-CoV-2 infections in infants. Sci Transl Med. 2026;18(838):eaea7097. Epub 20260225. doi: 10.1126/scitranslmed.aea7097. PubMed PMID: 41739901; PubMed Central PMCID: PMCPMC13317485.

45. Gonzalez-Kozlova E, Huang HH, Jagede OA, Tuballes K, Del Valle DM, Kelly G, et al. Tumor-Immune Signatures of Treatment Resistance to Brentuximab Vedotin with Ipilimumab and/or Nivolumab in Hodgkin Lymphoma. Cancer Res Commun. 2024;4(7):1726–37. doi: 10.1158/2767-9764.CRC-24-0252. PubMed PMID: 38934093; PubMed Central PMCID: PMCPMC11247952.

46. Li H, Zhang Y, Shang Y, Dai J, Ji S, Wu K. Identification of Treatment Targets in Allergic Conjunctivitis Through Proteome-Scale Mendelian Randomization Analysis. Mediators Inflamm. 2026;2026:6432686. Epub 20260131. doi: 10.1155/mi/6432686. PubMed PMID: 41624376; PubMed Central PMCID: PMCPMC12860419.

47. Valli A, Kuuliala K, Virtanen A, Kuuliala A, Palmroth M, Peltomaa R, et al. Tofacitinib treatment modulates the levels of several inflammation-related plasma proteins in rheumatoid arthritis and baseline levels of soluble biomarkers associate with the treatment response. Clin Exp Immunol. 2022;210(2):141–50. doi: 10.1093/cei/uxac085. PubMed PMID: 36124688; PubMed Central PMCID: PMCPMC9750823.

48. Zhang Y, Matzaraki V, Vadaq N, Blaauw MJT, Vos W, Groenendijk A, et al. Opposite effects of chronic HIV infection and antiretroviral medication on organismal and organ-specific biological aging. Nature Communications. 2026;17(1):2572. doi: 10.1038/s41467-026-69412-1.

49. Zhao H, Da F, Hou H, Chen X, Ling X, Wu S, et al. Metabolic dysbiosis as a predictor of immunological non-response among people living with HIV. Front Cell Infect Microbiol. 2026;16:1785624. Epub 20260515. doi: 10.3389/fcimb.2026.1785624. PubMed PMID: 42221578; PubMed Central PMCID: PMCPMC13219330.

50. Vadaq N, Groenendijk AL, dos Santos JC, Mehta K, Wit FWNM, Vos WAJW, et al. Longitudinal plasma proteomic signatures of elite and viremic spontaneous HIV controllers. Nature Communications. 2026;17(1):1173. doi: 10.1038/s41467-025-67939-3.

51. Hailu A, Musa A, Wasunna M, Balasegaram M, Yifru S, Mengistu G, et al. Geographical variation in the response of visceral leishmaniasis to paromomycin in East Africa: a multicentre, open-label, randomized trial. PLoS Negl Trop Dis. 2010;4(10):e709. Epub 20101026. doi: 10.1371/journal.pntd.0000709. PubMed PMID: 21049059; PubMed Central PMCID: PMCPMC2964287.

52. Khalil EA, Weldegebreal T, Younis BM, Omollo R, Musa AM, Hailu W, et al. Safety and efficacy of single dose versus multiple doses of AmBisome for treatment of visceral leishmaniasis in eastern Africa: a randomised trial. PLoS Negl Trop Dis. 2014;8(1):e2613. Epub 20140116. doi: 10.1371/journal.pntd.0002613. PubMed PMID: 24454970; PubMed Central PMCID: PMCPMC3894173.

53. Pasyar N, Alborzi A, Pouladfar GR. Evaluation of serum procalcitonin levels for diagnosis of secondary bacterial infections in visceral leishmaniasis patients. Am J Trop Med Hyg. 2012;86(1):119–21. doi: 10.4269/ajtmh.2012.11-0079. PubMed PMID: 22232461; PubMed Central PMCID: PMCPMC3247119.

54. Ahmed AA, Ahmed M, Theodorsson E, Nordlind K. Decreased concentrations of CGRP in Leishmania major murine cutaneous leishmaniasis. Neurosci Lett. 1998;246(3):149–52. doi: 10.1016/s0304-3940(98)00236-5. PubMed PMID: 9792614.

55. Ahmed AA, Wahbi AH, Nordlin K. Neuropeptides modulate a murine monocyte/macrophage cell line capacity for phagocytosis and killing of Leishmania major parasites. Immunopharmacol Immunotoxicol. 2001;23(3):397–409. doi: 10.1081/iph-100107339. PubMed PMID: 11694030.

56. Qiao X, Guo S, Meng Z, Gan H, Wu Z, Sun Y, et al. Advances in the study of death receptor 5. Front Pharmacol. 2025;16:1549808. Epub 20250312. doi: 10.3389/fphar.2025.1549808. PubMed PMID: 40144653; PubMed Central PMCID: PMCPMC11936945.

57. Forero-Torres A, Infante JR, Waterhouse D, Wong L, Vickers S, Arrowsmith E, et al. Phase 2, multicenter, open-label study of tigatuzumab (CS-1008), a humanized monoclonal antibody targeting death receptor 5, in combination with gemcitabine in chemotherapy-naive patients with unresectable or metastatic pancreatic cancer. Cancer Med. 2013;2(6):925–32. Epub 20131025. doi: 10.1002/cam4.137. PubMed PMID: 24403266; PubMed Central PMCID: PMCPMC3892397.

58. Fabie A, Mai LT, Dagenais-Lussier X, Hammami A, van Grevenynghe J, Stager S. IRF-5 Promotes Cell Death in CD4 T Cells during Chronic Infection. Cell Rep. 2018;24(5):1163–75. doi: 10.1016/j.celrep.2018.06.107. PubMed PMID: 30067973.

59. Salih MA, Ibrahim ME, Blackwell JM, Miller EN, Khalil EA, ElHassan AM, et al. IFNG and IFNGR1 gene polymorphisms and susceptibility to post-kala-azar dermal leishmaniasis in Sudan. Genes Immun. 2007;8(1):75–8. Epub 20061130. doi: 10.1038/sj.gene.6364353. PubMed PMID: 17136124; PubMed Central PMCID: PMCPMC2330095.

